# Proteomic-based Aging Clocks and MRI Markers of Cerebral Small Vessel Disease: ARIC and MESA

**DOI:** 10.64898/2026.04.02.26350087

**Authors:** Saeun Park, Shuo Wang, Jialing Liu, Timothy M Hughes, Erika P. Raven, Jelle Veraart, Mohamad Habes, Ruth Dubin, Rajat Deo, Wendy S. Post, Jerome I. Rotter, Alexis C. Wood, Peter Ganz, Behnam Sabayan, Weihong Tang, Josef Coresh, James S. Pankow, Keenan A. Walker, Pamela L. Lutsey, Weihua Guan, Anna E. Prizment, Sanaz Sedaghat

## Abstract

**Background:** This study investigates whether proteomic aging clocks (PACs) are associated with cerebral small vessel disease (CSVD).

**Methods:** We included participants from two US community-based cohorts: the Atherosclerosis Risk in Communities (ARIC) Study and the Multi-Ethnic Study of Atherosclerosis (MESA) Study. These analyses leveraged PACs that were developed in ARIC using proteomics measured by SomaScan in midlife (Visit 2; mean age 56 y; n=1,486) and late-life (Visit 5; mean age 76 y; n=1,496), trained on chronological age. Proteomic age acceleration (PAA) was calculated as residuals from regressing PACs on chronological age. 3T brain MRI data were collected in late-life. We examined associations of PAA with log-transformed white matter hyperintensity (WMH) volume using linear regression and with the presence of microbleeds, and subcortical, lacunar, and cortical infarcts using logistic regression. Associations of PACs with WMH volume and microbleeds were tested in MESA using proteins measured at Exam 1 (mean age 57 y; n=932) and Exam 5 (mean age 66 y; n=934). All associations were quantified per 5-year increase in PAA. All models were adjusted for demographics and cardiovascular risk factors.

**Results:** In ARIC, higher midlife PAA was associated with greater WMH volume (percent difference: 25% [95% CI: 13%, 39%]) and higher odds of subcortical infarcts (OR: 1.24 [1.02, 1.51]). Late-life PAA was associated with all CSVD markers: WMH volume (percent difference: 20% [8%, 34%]), cerebral microbleeds (OR: 1.40 [1.15, 1.69]), subcortical (OR: 1.80 [1.47, 2.22]), lacunar (OR: 1.80 [1.46, 2.23]), and cortical infarcts (OR: 1.39 [1.07, 1.82]). In MESA, higher late-life PAA was associated with greater WMH volume (28% [3%, 58%]) but not with microbleeds.

**Conclusion:** Accelerated proteomic aging is associated with a higher prevalence of MRI markers of CSVD, most predominantly in late-life. Understanding this relationship may help stratify those at higher risk of CSVD at an early stage.

## Introduction

Cerebral small vessel disease (CSVD) represents a range of pathological abnormalities in the brain’s microvasculature including white matter hyperintensities (WMHs), lacunar infarcts, and microbleeds (1–3). CSVD may remain subclinical for years. However, a high burden of CSVD or coexistence of different pathologies are strongly associated with increased risk of depression, cognitive decline, incident dementia, and stroke (3). CSVD contributes to ∼25% of ischemic strokes and ∼50% of all dementia cases (3, 4). It is the most common pathology underlying vascular cognitive impairment and vascular dementia, and its presence significantly increases the risk of existing subclinical neurodegenerative pathologies developing into dementia (5).

Although chronological age is the most significant determinant of CSVD, it may be a limited predictor of CSVD risk, as individuals of the same chronological age can have different biological aging (6, 7). Biological aging clocks, defined as cellular or molecular changes associated with aging, have been emerging as promising tools to track individuals’ biological aging trajectories and predict age-related adverse health outcomes (8). Proteomic aging clocks (PACs) have shown potential to estimate biological age and stratify risk for age-related diseases, with strong associations observed for all-cause mortality risk and age-related diseases including Alzheimer’s disease and related dementias (9–11).

Previously, we developed PACs in dementia-free participants from the Atherosclerosis Risk in Communities (ARIC) study, using SomaScan-measured proteomics from plasma samples collected at midlife (ages 46–70, 1990–92, Visit 2) and late-life (ages 66–90, 2011–13, Visit 5). PACs were constructed separately for each life stage by training proteins against chronological age (10, 11). In our recent work, we confirmed that PACs at both life stages were strongly associated with dementia risk (11). Given that CSVD is a major contributor to cognitive impairment and dementia, particularly vascular dementia, it is plausible that PACs are also associated with a higher burden of CSVD. However, the relationship between PACs and the prevalence of CSVD has not yet been established. Therefore, in this study, we examined whether proteomic age acceleration (PAA) (defined as the deviation of biological age from chronological age based on midlife or late-life PACs) was associated with the prevalence of late-life CSVD, including WMH volume, cerebral microbleeds, and subcortical, lacunar, and cortical infarcts. We hypothesized that PAA at both life stages is associated with greater WMH volume and a higher prevalence of cerebral microbleeds and different types of infarcts.

This study was conducted in two large population-based cohorts. First, PACs were tested in ARIC, a prospective study of mostly White and Black men and women with proteomic data measured both 20 years before and at the time of brain MRI scanning. Then, the findings were externally validated in the Multi-Ethnic Study of Atherosclerosis (MESA), another prospective cohort including ethnically diverse participants self-identifying as Black, White, Chinese, and Hispanic.

## Methods ARIC study

### Study population

This study included White and Black male and female participants of the ongoing ARIC study. The ARIC study is a large, prospective cohort study to investigate the cause and clinical outcomes of atherosclerosis among U.S. adults (12). At Visit 1 (1987–89), ARIC enrolled 15,792 participants aged 44 to 66 years from four U.S. communities: Washington County, MD; Forsyth County, NC; Jackson, MS; and suburban Minneapolis, MN. So far, ten follow-up examinations have been completed including Visit 2 (1990–92) and Visit 5 (2011–13) that are relevant to the present analysis. Of the 6,538 participants who attended Visit 5, a subset of participants (N = 3,180) was invited for brain MRI scanning as part of the ARIC Neurocognitive Study (NCS) (13). These participants consisted of three groups: individuals who participated in the ARIC Brain MRI study in 2004–2006, those with evidence of cognitive impairment at Visit 5, and an age- and field center–stratified random sample of cognitively healthy participants (7). Of Visit 2 and Visit 5 participants, 1,948 and 1,972, respectively, completed brain MRI scanning. Among participants who attended Visit 2 (midlife) and Visit 5 (late-life), we excluded individuals who did not participate in the ARIC-NCS brain MRI scan; those with missing brain MRI data (based on missingness in lacunar infarct); those without protein data; those with prevalent dementia at the corresponding visit; and participants who self-identified as a race other than White or Black, as well as Black participants from the Minneapolis or Washington County field centers due to small sample sizes (**Figure S1** and **S2**). After exclusions, the analytic samples included 1,486 midlife participants and 1,496 late-life participants.

### Measurement

#### Proteomics measurement (Visit 2 and Visit 5)

In ARIC, 5,284 aptamers were measured by a SOMAmer (Slow Off-rate Modified Aptamer)-based assay called SomaScan (v4.0) (SomaLogic, Inc., USA) in stored frozen plasma samples collected at midlife and late-life (14). The SomaScan assay uses a multiplexed, modified DNA-based aptamer technology to measure protein concentrations (15). The details of SomaScan assay and data normalization process have been described previously (14, 15). Among the measured aptamers, those with a Bland-Altman coefficient of variation (CVBA) greater than 50% or a variance of less than 0.01 on the log scale, or binding to mouse Fc-fusion, contaminants, or non-proteins were excluded (16). After the exclusion, 4,955 aptamers (4,712 unique proteins) were included in midlife and late-life. Protein concentrations were reported in relative fluorescence units (RFU) and log2-transformed to adjust for skewedness (10).

### MRI Markers of Cerebral Small Vessel Disease (Visit 5)

Details of brain MRI scanning and interpretation have been reported previously (7). Briefly, the ARIC Visit 5 (2011–13) brain MRI scans were performed by using 3T brain scanner at each study site (Maryland: Siemens Verio; North Carolina: Siemens Skyra; Minnesota: Siemens Trio; Mississippi: Siemens Skyra) based on the identical protocols. The following sequences were performed on all participants: Magnetization prepared - rapid gradient echo (MP-RAGE) (1.2-mm slices), T2-weighted gradient-recalled echo (T2*GRE) (4-mm slices), and axial T2 fluid-attenuated inversion recovery (FLAIR) (5-mm slices) (17). WMH volume and infarcts were measured using a T2-weighted FLAIR imaging and microbleeds were assessed based on T2*GRE sequences (17). WMH volume was quantified using a semiautomated segmentation algorithm on T2-weighted FLAIR images and recorded in cubic centimeter (18–20). Microbleeds were defined based on T2*GRE sequences of 5 mm in maximum diameter and were categorized into lobar (cortical gray and subcortical or periventricular white matter), deep (deep gray matter, basal ganglia, thalamus, and the white matter of the corpus callosum, internal, external, and extreme capsule), and infratentorial microbleeds (brain stem and cerebellum) (17, 21). Cortical infarcts were defined as hyperintense T2 FLAIR lesions with a minimum extent >10 mm, extending to the cortical surface that includes cortical gray matter and may include underlying white matter (13). Lacunar infarcts were defined as subcortical T2 FLAIR lesions with a hyperintense rim and a dark center, ranging from 3 to 20 mm in size, located in the basal ganglia, thalamus, brain stem, internal capsule, deep cerebellum, and subcortical white matter (22).

Subcortical infarcts were assessed as higher intense T2 FLAIR lesions with a dark center with a minimum of 3mm in diameter in the white matter, infratentorial, and central gray/capsular regions, and distinguishable from perivascular spaces (13). Both cerebral microbleeds and infarcts were assessed by a trained imaging technician and confirmed by radiologists (18, 19). Total intracranial volume was measured on MP-RAGE sequences using image analysis software (FreeSurfer; http://surfer.nmr.mgh.harvard.edu) (18).

### Other variables (covariates or exclusion criteria variables)

Sex, race (Black or White), and educational attainment (less than high school, high school equivalent, or at least some college degree) were self-repfforted using a questionnaire at ARIC Visit 1. Other covariates were measured at both ARIC Visits 2 and 5. Smoking status (current, former, or never users) was self-reported. Body mass index (BMI) was computed as weight divided by height squared (kg/m^2^). Diabetes was defined as a self-reported history of physician- diagnosed diabetes, use of diabetes medication within the past two weeks, a fasting blood glucose level ≥ 126 mg/dL, a non-fasting blood glucose level ≥ 200 mg/dL, or a hemoglobin A1C level ≥ 6.5% (23). Hypertension was identified by a systolic blood pressure 140 mm Hg or higher, a diastolic blood pressure 90 mm Hg or higher, or the use of antihypertensive medications (24). Plasma total cholesterol levels were quantified employing enzymatic methods (25).

Estimated glomerular filtration rate (eGFR) was calculated using the race-free Chronic Kidney Disease Epidemiology Collaboration (CKD-EPI) 2021 creatinine and cystatin-C equation (26). The *APOE* genotypes were assessed using TaqMan assays and the ABI 7700 Sequence Detection System (Applied Biosystems, Foster City, CA) (27). Prevalent dementia was defined as dementia diagnosed before or at the ARIC follow-up visit (V2 for midlife and V5 for late-life). Briefly, dementia cases were ascertained using three sources of information: (1) in-person cognitive testing during follow-up examinations and an informant interview, (2) annual telephone interviews or an informant telephone interview, and (3) hospitalization and death certificate codes. Additional details on dementia ascertainment have been published elsewhere (28).

Prevalent stroke was defined as any stroke event identified before or at the ARIC follow-up visit, based on information from follow-up examinations, annual telephone interviews, and hospitalization record (29).

### MESA study

#### Study population

We included men and women participants of the Multi-Ethnic Study of Atherosclerosis (MESA) study to replicate the association between PAC and CSVD in ARIC. MESA is an ethnically diverse, community-based, prospective cohort study that was launched in 2000 to investigate the risk factors influencing the transition of subclinical cardiovascular disease to overt cardiovascular events. At baseline (Exam 1, 2000-02), MESA enrolled 6,814 participants aged 45-84 free from cardiovascular diseases. As part of the screening form, participants self-identified their race and ethnicity, resulting in recruitment of 38% White, 28% Black, 22% Hispanic, and 12% Chinese American participants from six U.S. field centers: Baltimore City and Baltimore County, MD; Chicago, IL; Forsyth County, NC; Los Angeles County, CA; New York, NY; and St. Paul, MN. Since the baseline, six follow-up examinations have been conducted, of which Exam 1 (2002–04) and Exam 5 (2010–11) are included in the present study (30). To validate the associations between PACs and brain MRI markers of CSVD observed in ARIC, we conducted replication analyses in MESA using Exam 1 (ages 45–84) and Exam 5 (ages 55–94). These exams were selected because their age ranges comparable to ARIC midlife (46–68 years) and late-life (67–90 years) visits, and proteomics data were available at both time points. In MESA, participants with clinically recognized atrial fibrillation were oversampled for the Atrial Fibrillation ancillary study at Exam 6 (2016–2018). Among ancillary study participants, a subset who were eligible (e.g., without metal implants) and willing completed brain MRI scanning, which were conducted from March 2018 through August 2019 (31, 32). Among MESA Exam 1 or 5 participants, 1,057 completed their brain MRI scan as part of the Atrial Fibrillation ancillary study. At both exams, we excluded participants without protein data, those who did not undergo brain MRI scanning, and those with prevalent dementia at the corresponding exam (**Figure S3** and **S4**). The final analytic samples included 932 participants at Exam 1 and 934 at Exam 5.

## Measurement

### Proteomics

At MESA Exam 1 and 5, 7,329 aptamers (corresponding to 6,430 proteins) were quantified at the SomaLogic using EDTA plasma samples collected by a standardized protocol at each visit and stored at −80°C. Proteomics were measured using SomaScan v4.1 assay, which covers all the aptamers measured by the earlier v4.0 assay used in ARIC (33). The means and distributions of randomly selected aptamers measured in ARIC and MESA were similar (11). The laboratory used standard SomaLogic quality control methods for normalization and calibration (34). Protein values, measured in RFU, were transformed into log2 scale for normalization and outliers >5 standard deviations from the sample mean on the log_2_ scale were winsorized (35).

### Brain MRI markers

Brain MRI scanning was performed as previously described (36) as part of the Atrial Fibrillation ancillary study a median of 18 months (IQR, 16–20 months) after Exam 6 (2018–2019) using 3T brain scanner at each study site (University of California Los Angeles, Columbia University, John Hopkins University, Northwestern University, University of Minnesota: Prisma VE11C; University of California Los Angeles, Wake Forest University: Skyra VD11B) based on the identical protocols. Structural MRI brain sequences included T1-weighted, T2-weighted, T2 FLAIR, and a susceptibility-weighted imaging (SWI) quantitative susceptibility mapping (QSM) sequence. WMH volume was quantified by inhomogeneity-corrected and co-registered FLAIR and T1-weighted images applying a deep learning-based segmentation method (37). Microbleeds were initially identified by a deep learning-based method that used T2-weighted, quantitative susceptibility mapping (QSM) and susceptibility-weighted imaging (SWI) (38). Identified lesions were then reviewed by a radiologist, who made the final classification. and grouped them into lobar (frontal, parietal, temporal, occipital, and insula), deep (basal ganglia, thalamus, internal capsule, corpus callosum, and deep and periventricular white matter), and infratentorial (brainstem and cerebellum). MRI markers for infarcts (subcortical, lacunar, and cortical) that were available in ARIC were not measured in MESA.

### Other variables

Race/ethnicity (Black, White, Chinese, and Hispanic) were collected as part of the MESA screening protocol, and sex, race/ethnicity and educational attainment was self-reported by questionnaire at Exam1. Educational attainment was categorized into less than high school, high school equivalent, and at least some college degree. Other covariates were collected at both exams. Smoking status (never, former, and current smoker) was self-reported. Body mass index (BMI) was measured by trained study personnel at the clinic exam and calculated as weight divided by height squared (kg/m^2^). Diabetes was defined as fasting glucose > 6.99 mmol/L (126 mg/dL), self-report of a prior diagnosis, or self-reported use of hypoglycemic medication. Total cholesterol was measured using the cholesterol oxidase method (Roche Diagnostic, Indianapolis, IN) (39). Hypertension was defined as the use of hypertension medication, a diagnosis of hypertension, or SBP ≥ 140 mm Hg or DBP ≥ 90 mm Hg. eGFR was calculated using the creatinine and cystatin-C based CKD-EPI equation (40). *APOE* isoforms were estimated from single nucleotide polymorphisms rs429358 and rs741 using the algorithm described elsewhere (41). Prevalent dementia was defined as dementia cases ascertained based on hospitalization codes recorded before or at the study examination. Detailed information on dementia ascertainment has been described previously (42). Prevalent stroke was defined as any stroke event that occurred before or at the time of the exam, identified through phone interviews or medical records for hospitalizations or outpatient diagnoses. Details on methods applied to ascertain stroke events were reported previously (43).

## Statistical Analysis

### Development of Proteomic-based aging clocks (PACs) in ARIC

Details on the creation of PACs are available elsewhere (10, 11). Briefly, we previously created dementia-free PACs separately at ARIC midlife and late-life visits. To construct the PAC at each life stage, we randomly selected two-thirds of the participants who did not develop dementia until 2019 (by the end of Visit 7) and used them as the training set. The remaining one-third of participants, who were also dementia-free until 2019, served as the test set at the corresponding life stages. In the training set, we applied elastic net regression to train the ARIC dementia-free PACs against chronological age and obtain the weight (regression coefficient) for each aptamer. Lambda value in the elastic net model was selected based on 10-fold cross-validation. Using the proteins and weights determined in the training set, we computed PACs for all dementia-free participants from the pooled training and test sets as a weighted sum of aptamers using the following formula: 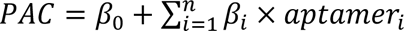, where *aptamer_i_* is the level of the *ith* aptamer (10, 11). This resulted in the selection of 1,176 aptamers (1,160 proteins) in ARIC midlife and 618 aptamers (613 proteins) in ARIC late-life participants (**Table S1** and **S2**). We computed PAA as the residuals from regressing PAC on chronological age to assess the effect of PACs independent of age. A positive value of PAA indicates that the proteomic biological age is older than the person’s chronological age.

### Proteomics age acceleration and MRI markers of CSVD in ARIC

To examine associations of PAA (per 5-year increase) with CSVD, we used linear regression models for natural log-transformed WMH volume and logistic regression models for the presence of cerebral microbleeds and different types of infarcts (subcortical, lacunar, and cortical). Estimates from the log-linear regression were reported as the percent difference in geometric mean WMH volume per 5-year increase in PAA, obtained by exponentiating the regression coefficients. All models were adjusted for chronological age, sex, a combined race and center variable, education attainment, BMI, smoking status, eGFR, diabetes, and total cholesterol levels. The combined variable for race and center included five groups: Black-Mississippi, Black-North Carolina, White-North Carolina, White-Maryland, and White-Minnesota participants. This approach is widely used in ARIC cohort analyses due to the recruitment strategies adopted. The Jackson study site recruited only Black adults, while the remaining three study sites recruited participants using probability sampling, which resulted in participants being almost exclusively White in the Minnesota and Washington County field sites, and a mix of White and Black participants in North Carolina (18). Models using WMH volume as the outcome were further adjusted for total intracranial volume. In all analyses, we applied inverse sampling weights to adjust for potential selection bias associated with the stratified random sampling that was used to select participation into the brain MRI study and the probability of attrition (i.e., failure to complete the Visit 5 MRI study due to nonattendance or death) and to represent the full ARIC visit 5 cohort (18). Sampling weights were computed as the product of the inverse sampling fraction and the inverse probability of exam completion to account for being unwilling or unable to attend visits (13).

### External validation in MESA

Associations between midlife and late-life PACs with MRI markers of CSVD were replicated in MESA Exam 1 and Exam 5, respectively. In MESA, PACs were replicated by applying the weights calculated in ARIC for each life stage to the protein concentrations measured at MESA Exam 1 (for midlife) and Exam 5 (for late-life). We then computed PAA from the replicated PACs and tested its association with MRI markers, using linear regression for natural log-transformed WMH volume and logistic regression for cerebral microbleeds (presence vs. absence) at each exam. All models were adjusted for the same set of covariates as in ARIC: age, sex, race, center, educational attainment, BMI, smoking status, eGFR, diabetes, and total cholesterol. Consistent with the analyses in ARIC, the models for WMH volume were further adjusted for total intracranial volume.

### Sensitivity analyses in ARIC and MESA

In ARIC, to explore the factors driving the differences in the associations between PACs at each life stage and brain MRI markers, we constructed a modified version of the midlife PAC using the weights calculated for the 618 aptamers (613 proteins) included in the late-life PAC, while keeping other factors constant (i.e., using midlife participants and midlife protein values).

Similarly, we created modified late-life PAC by replacing the weights with those calculated for the 1,176 aptamers (1,160 proteins) included in the midlife PAC. We then computed PAA based on the reconstructed PACs at each time point and examined their associations with brain MRI markers of CSVD.

In both ARIC and MESA, to explore whether participants’ stroke history influenced the results, we repeated the analyses on PAA and brain MRI markers of CSVD after excluding individuals who had experienced a clinical stroke before or during each life stage. We also examined the association between PAA and microbleeds by location using multinomial logistic regression after further categorizing them as none, strictly lobar (microbleeds in lobar regions only), strictly deep (microbleeds in deep or infratentorial regions without lobar microbleeds), or mixed (microbleeds in deep/infratentorial regions with lobar microbleeds). Multinomial logistic regression estimates were reported as relative odds ratios based on exponentiated effect estimates from the model.

Lastly, for the significant associations found in the main analysis, we assessed whether they changed by *APOE* ε4 carriership (no ε4 alleles vs. one or two ε4 alleles), sex, and race using stratified analyses.

## Results

### Baseline characteristics of participants in ARIC and MESA

The baseline characteristics of midlife and late-life participants in the ARIC and MESA cohorts are summarized in **Table 1**. In ARIC, the mean ± SD age of participants was 56 ± 5 years in midlife and 76 ± 5 years in late-life. At both life stages, approximately 60% of participants were women, and 25% were Black. In MESA, the mean age of midlife participants was 57 ± 8 years, whereas that of late-life participants was 66 ± 8 years. In MESA, at both life stages, 52% were women, and the largest proportion was White (41%), followed by Black (25%) and Hispanic (19%). In both ARIC and MESA, the proportion of current smokers was higher in midlife than late-life (ARIC: 16% vs 5%; MESA: 11% vs 7%), while hypertension and diabetes were more prevalent in late-life (ARIC: 25% vs 74% and 8% vs 33%; MESA: 29% vs 50% and 18% vs 36%). Median WMH volume was 12 mL in ARIC and 2.8 mL in MESA, and microbleeds were present in approximately 24% of ARIC and 34% of MESA participants. In ARIC, PAA ranged from −7.13 to 8.04 in midlife and from −5.96 to 13.85 in late-life; in MESA, it ranged from −7.25 to 8.80 in midlife and from −5.54 to 6.91 in late-life (**Table S3**). The correlations between chronological age and midlife or late-life PACs were strong in both cohorts (all r > 0.83).

**Table 1.** Baseline characteristics of midlife (ARIC Visit 2: 1990–92; MESA Exam 1: 2000–02) and late-life participants (ARIC Visit 5: 2011–13; MESA Exam 5: 2010–11) – ARIC and MESA.

| Characteristics | ARIC* |  | MESA† |  |
| --- | --- | --- | --- | --- |
|  | Midlife<br>N=1,486 | Late-life<br>N=1,496 | Midlife<br>N=932 | Late-life<br>N=934 |
| **Demographics** |  |  |  |  |
| Mean age, years (SD) | 55.7 (5.2) | 76.2 (5.2) | 56.7 (8.3) | 66.26 (8.23) |
| Age range, years | 46–68 | 67–90 | 44–81 | 54–91 |
| Gender, N (Female %) | 872 (58.7) | 879 (58.8) | 563 (53.3) | 499 (53.4) |
| Race, N (Black %) |  |  |  |  |
| Black | 369 (24.8) | 367 (24.5) | 235 (25.2) | 235 (25.2) |
| White | 1,117 (75.2) | 1,129 (75.5) | 378 (40.6) | 381 (40.8) |
| Hispanic/Latino | - | - | 178 (19.1) | 175 (18.7) |
| Chinese | - | - | 141 (15.1) | 143 (15.3) |
| Education, N (%) |  |  |  |  |
| Less than High School | 193 (13.0) | 191 (12.8) | 98 (10.5) | 99 (10.6) |
| High School Equivalent | 634 (42.7) | 627 (41.9) | 142 (15.2) | 142 (15.2) |
| At least some college | 657 (44.3) | 677 (45.3) | 692 (74.2) | 693 (74.2) |
| **Lifestyle/Comorbidity Factors** |  |  |  |  |
| Mean BMI, kg/m<sup>2</sup> (SD) | 27.5 (4.9) | 28.6 (5.6) | 27.7 (5.1) | 28.1 (5.2) |
| Smoking Status, N (%) |  |  |  |  |
| Current Smoker | 231 (15.6) | 74 (5.2) | 104 (11.2) | 67 (7.2) |
| Former Smoker | 553 (37.3) | 715 (50.7) | 343 (36.9) | 432 (46.3) |
| Never Smoked | 699 (47.1) | 622 (44.1) | 483 (51.9) | 434 (46.5) |
| Hypertension, N (%) | 367 (24.8) | 1,100 (74.2) | 277 (29.7) | 466 (49.9) |
| Diabetes, N (%) | 124 (8.4) | 490 (33.0) | 173 (18.6) | 339 (36.3) |
| Mean eGFR, ml/min/1.73 m<sup>2</sup> (SD) | 101.3(14.1) | 68.9 (18.3) | 80.9 (14.2) | 70.9 (14.5) |
| Mean Cholesterol, mg/dL (SD) | 208.9 (36.9) | 182.3 (41.6) | 194.43 (33.05) | 185.7 (36.9) |
| History of stroke, N (%) | 11 (0.7) | 49 (3.3) | 0 (0.0) | 10 (1.1) |
| **Brain MRI markers‡** |  |  |  |  |
| Mean age at brain MRI measurement, years (SD) | 76.4 (5.3) | 76.2 (5.2) | 74.0 (8.2) | 74.0 (8.2) |
| Median WMH volume, mL (Q1, Q3) | 12.0 (6.7, 21.9) | 11.2 (6.3, 20.6) | 2.8 (1.2, 7.5) | 2.8 (1.2, 7.5) |
| Median intracranial volume, mL (Q1, Q3) | 1372.6 (1271.9, 1491.9) | 1368.4 (1268.2, 1491.0) | 1355.4 (1251.5, 1452.8) | 1355.8 (1251.6, 1454.5) |
| Presence of cerebral microbleeds, N (%) | 361 (24.5) | 355 (23.9) | 299 (34.7) | 294 (34.1) |
| Presence of subcortical infarcts, N (%) | 287 (19.3) | 282 (18.9) | - | - |
| Presence of lacunar infarcts, N (%) | 270 (18.2) | 264 (17.6) | - | - |
| Presence of cortical infarcts, N (%) | 161 (10.8) | 151 (10.1) | - | - |

Abbreviations: ARIC = Atherosclerosis Risk in Communities, MESA = Multi-Ethnic Study of Atherosclerosis, SD = standard deviation, BMI = body mass index, eGFR = estimated glomerular filtration rate, MRI = magnetic resonance imaging, Q1 = first quartile, Q3 = third quartile, WMH = white matter hyperintensity.
\* Missing values (n) in ARIC: education (Visit 2: n=2; Visit 5: n=1); BMI (Visit 5: n=6); smoking status (Visit 2: n=3; Visit 5: n=85); hypertension (Visit 2: n=6; Visit 5: n=13); diabetes (Visit 2: n=2; Visit 5: n=12); eGFR (Visit 2: n=41; Visit 5: n=2); history of stroke (Visit 2: n=1); WMH volume (Visit 2: n=1; Visit 5: n=1); cerebral microbleeds (Visit 2: n=13; Visit 5: n=13).
† Missing values (n) in MESA: smoking status (Exam 1: n=2; Exam 5: n=1); eGFR (Exam 1: n=2); history of stroke (Exam 5: n=1); WMH volume (Exam 1: n=20; Exam 5: n=3); cerebral microbleeds (Exam 1: n=70; Exam 5: n=72).
‡ For each cohort, brain MRI markers were measured at a single time point: in ARIC at Visit 5 (2011–2013) and in MESA Atrial Fibrillation ancillary study (2018-19). Subcortical, lacunar, and cortical infarcts were assessed only in ARIC.

#### Association between proteomic age acceleration and brain MRI markers of CSVD in ARIC

Findings from the linear and logistic regression models examining the association between PAA in midlife or late-life with brain MRI markers of CSVD in late-life, adjusted for the full set of covariates, are presented in **Figure 1**. Each 5-year higher midlife PAA was prospectively associated with a 25% larger WMH volume (95% CI: 13%–39%) and higher odds of subcortical infarcts (OR = 1.24, 95% CI: 1.02–1.51) measured at late-life (approximately 21 years after the protein measurement at midlife), but no significant associations were found with other MRI markers of CSVD. On the other hand, late-life PAA was cross-sectionally associated with all five MRI markers. A 5-year higher late-life PAA was linked to a 20% greater WMH volume (95% CI: 8%–34%) and higher odds of cerebral microbleeds (OR = 1.40, 95% CI: 1.15–1.69), subcortical infarcts (OR = 1.80, 95% CI: 1.47–2.22), lacunar infarcts (OR = 1.80, 95% CI: 1.46–2.23), and cortical infarcts (OR = 1.39, 95% CI: 1.07–1.82). By microbleed anatomical location, each 5-year higher late-life PAA was associated with 1.73-fold higher odds of having strictly lobar microbleeds (95% CI: 1.25–2.40) and 2.22-fold higher odds of having mixed microbleeds (95% CI: 1.30–3.78), relative to having no microbleeds (**Table S4**).

**Figure 1.**
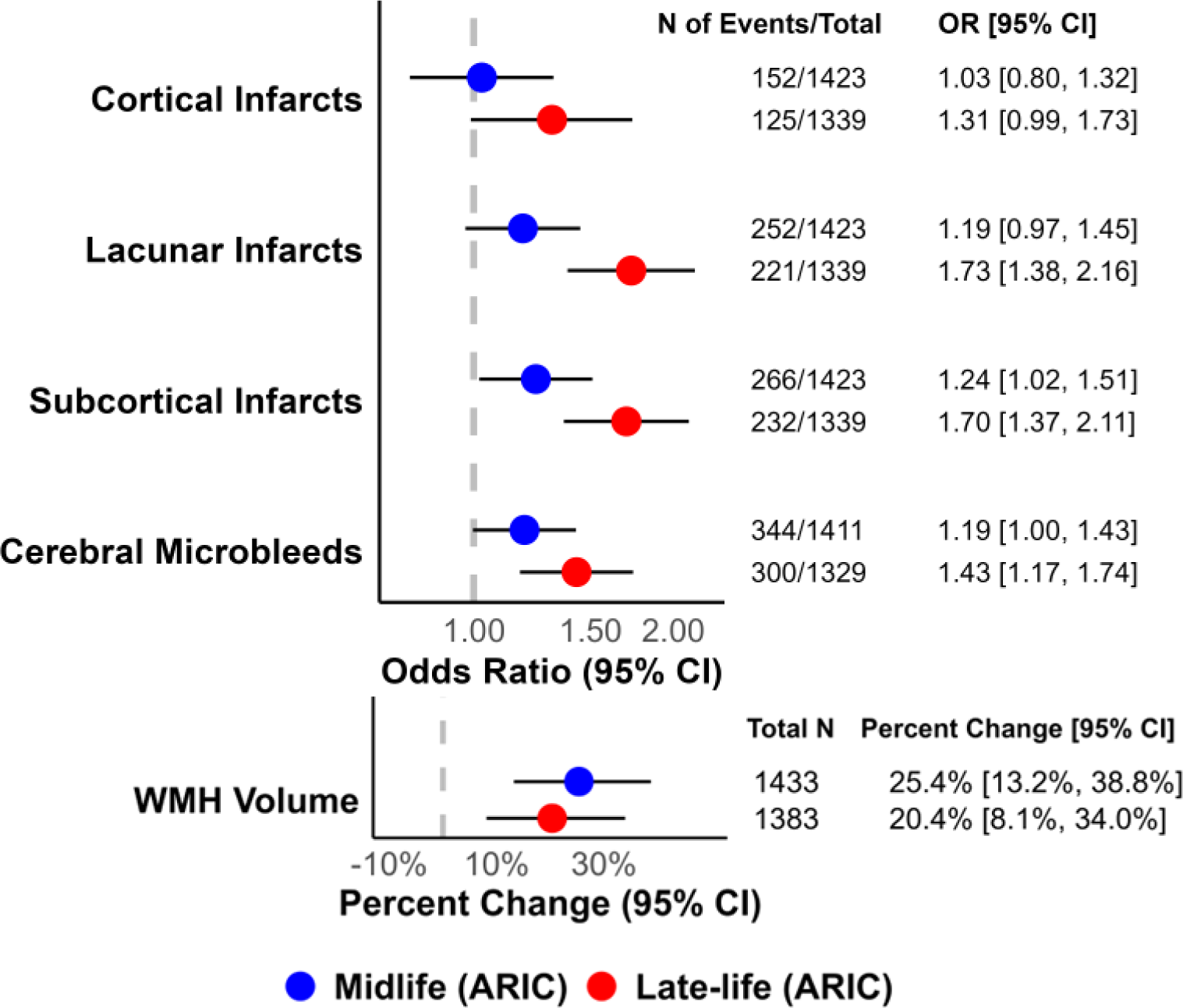
Associations of proteomic age acceleration, per 5-years, at midlife (Visit 2, 1990–92) and late-life (Visit 5, 2011–13) with magnetic resonance imaging (MRI) markers of cerebral small vessel disease in the ARIC Neurocognitive Study (Visit 5, 2011–13). The plot shows odds ratios for cortical infarcts, lacunar infarcts, subcortical infarcts, and cerebral microbleeds, as well as the percent change in white matter hyperintensity volume, per 5-year increase in proteomic age acceleration at midlife and late-life in ARIC. All models were adjusted for chronological age at mid/late-life, sex, race-center, education, BMI, smoking status, hypertension, diabetes status, cholesterol, and estimated glomerular filtration rate. Model for white matter hyperintensity volume were additionally adjusted for intracranial volume. Abbreviations: ARIC = Atherosclerosis Risk in Communities, PAC = proteomic-based aging clock, OR = odds ratio, WMH = white matter hyperintensity.

### External validation in MESA

In MESA, each 5-year higher late-life PAA, was prospectively associated with 28% larger WMH volume (95% CI, 4%–58%), measured approximately 8 years after the protein measurements (**Figure 2**) but midlife PAA was not significantly associated with WMH volume. At both stages of life, no significant associations were observed between PAA and microbleeds at different anatomical locations (strictly lobar vs. strictly deep vs. mixed) (**Table S4**).

**Figure 2.**
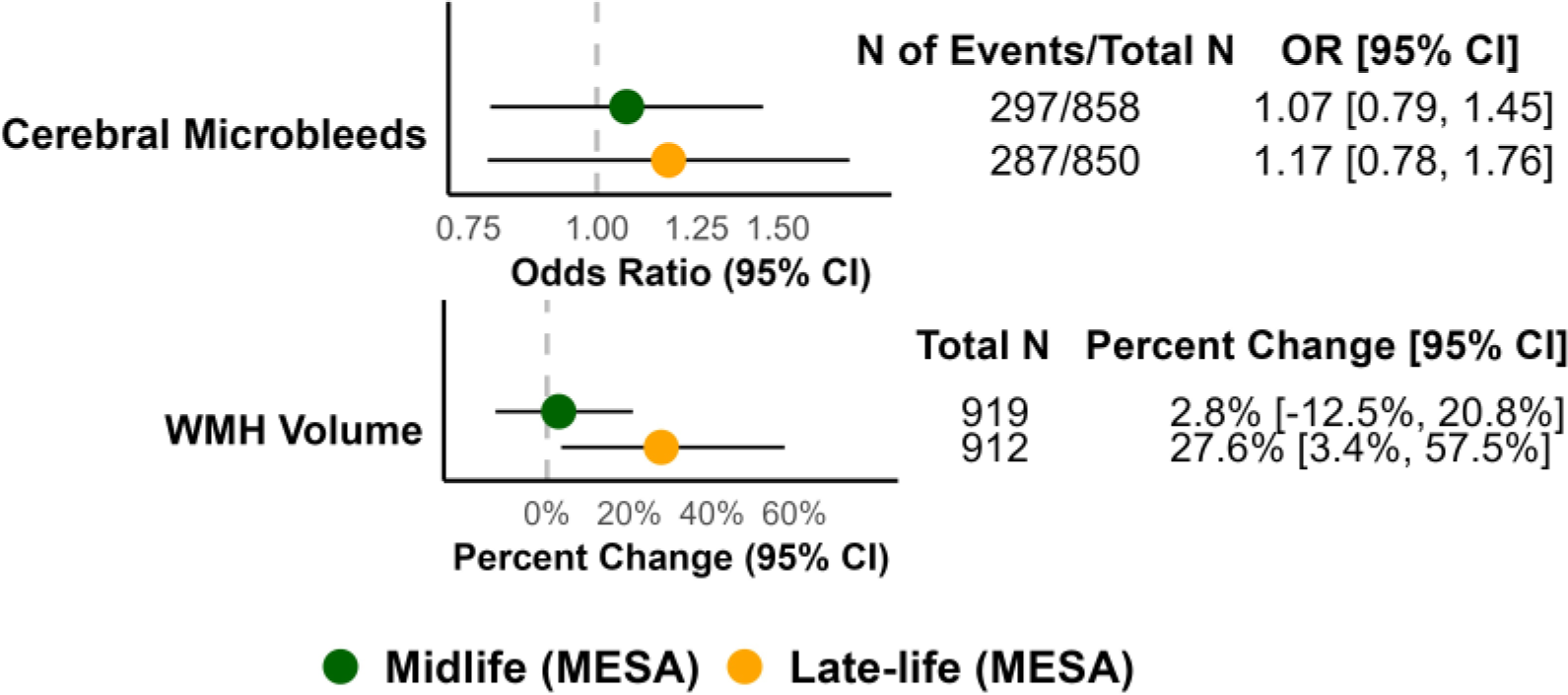
Associations between proteomic-based age acceleration, per 5-years, at midlife (Exam 1, 2000-02) and late-life (Exam 5, 2010–11) with magnetic resonance imaging (MRI) markers of cerebral small vessel disease in the MESA Atrial Fibrillation ancillary study (2018–19). The plot shows odds ratios for cerebral microbleeds and the percent change in white matter hyperintensity volume, per 5-year increase in proteomic age acceleration at midlife and late-life in MESA. All models were adjusted for chronological age at mid/late-life, sex, race-center, education, BMI, smoking status, hypertension, diabetes status, cholesterol, and estimated glomerular filtration rate. Model for white matter hyperintensity volume were additionally adjusted for intracranial volume. All models were adjusted for chronological age at mid/late-life, sex, race, center, education, BMI, smoking status, hypertension, diabetes status, cholesterol, and estimated glomerular filtration rate. Model for white matter hyperintensity volume were additionally adjusted for intracranial volume. Abbreviations: MESA = Multi-Ethnic Study of Atherosclerosis, PAC = proteomic-based aging clock, OR = odds ratio, WMH = white matter hyperintensity.

### Sensitivity analyses in ARIC and MESA

In ARIC, when we used midlife PAA based on midlife protein levels but with protein weights from the late-life PAC, its associations with MRI markers were attenuated compared with those for the original midlife PAA (**Figure S5**). Similarly, when we used late-life PAA computed by applying midlife PAC weights to late-life protein levels, the effect sizes for three of the five MRI markers were smaller than those for the original late-life PAA (**Figure S6**). This analysis was conducted only in ARIC to assess whether the associations were driven by model weights and protein selection or by protein levels.

In both the ARIC and MESA cohorts, the associations between midlife or late-life PAA and MRI markers remained consistent after excluding participants with prevalent stroke (**Figure S7** and **S8**). Stratified analyses provided some evidence of potential effect modification by *APOE* ε4 carriership, sex, and race (**Tables S5–S7**). For example, in ARIC, the association between late-life PAA and the presence of cortical infarcts was stronger among *APOE* ε4 carriers compared to non-carriers (p-interaction < 0.05). However, no consistent trend of effect modification was observed across MRI markers, life stages, or cohorts.

## Discussion

In a large community-based cohort study, we found that higher midlife PAA was associated with larger WMH volume and a higher prevalence of subcortical infarcts in late-life. Similarly, higher late-life PAA was associated with larger WMH volume and a higher prevalence of subcortical infarcts and was additionally associated with a higher prevalence of cerebral microbleeds and of lacunar and cortical infarcts. The association between late-life PAA and WMH volume was replicated in an external cohort. All associations were independent of chronological age, cardiovascular risk factors, and stroke history.

Several studies have investigated the potential link between epigenetics clocks based on DNA methylation (DNAm) and CSVD, but the findings remain inconclusive (6, 44–47). A few cross-sectional studies have reported that age acceleration, as measured by epigenetic clocks (e.g., HorvathAge, HannumAge, and GrimAge), is associated with a higher WMH burden (6, 45, 47). However, other cross-sectional studies have found negative or no associations between HorvathAge and WMH volume (44, 46). Two prior population-based cohort studies with large sample size have reported links between PACs and all-cause mortality and aging-related diseases including dementia and stroke (9, 11). Although the studies used different proteomic platforms (Olink vs SomaScan) with less than 50% overlap in the proteins included in the clocks, they consistently reported that having a higher proteomics-based biological age relative to chronological age was associated with a higher risk of dementia. Yet, whether PACs are associated with CSVD, a major contributor to both stroke and dementia, remained unknown. In the present study, in the ARIC cohort, we found that PAA was associated with brain MRI markers of CSVD. In the external MESA cohort, we validated the positive association between late-life PAA and WMH volume, but we did not find an association with the prevalence of microbleeds. Infarcts (subcortical, lacunar, and cortical) were not available in MESA, which precluded replication of the ARIC analyses.

Replication of the ARIC findings in MESA may have been limited for several reasons. First, the smaller sample size and broader age range in MESA may have reduced statistical power and limited our ability to detect associations. Second reason could be related to population differences included across two cohorts. Participants at MESA Exam 5 and at the time of MRI scanning (median, 18 months [IQR, 16–20 months] after Exam 6) were, on average, younger than ARIC participants at late-life, when the late-life clocks were developed and MRI scanning was performed. In addition, because MESA intentionally enrolled individuals free of clinical cardiovascular disease at baseline, MESA participants were likely healthier overall than ARIC participants at the time of MRI measurement. This may have reduced the burden and severity of neuroimaging abnormalities and limited the power to detect associations. Third, for microbleeds specifically, additional factors may have played a role when replicating the ARIC analyses in MESA. In MESA, participants with atrial fibrillation, who were likely to have a higher microbleed burden, were oversampled for the ancillary MRI study (32, 49). In addition, MESA used a deep learning–based method with higher sensitivity for detecting microbleeds than the approach used in ARIC (48). Together, these factors may have increased the observed microbleed burden in MESA relative to ARIC and reduced the comparability of microbleed findings across the two studies.

We found that PAA measured in late life was more strongly associated with markers of CSVD than PAA measured in midlife. The stronger association of late-life PAA and CSVD may be explained by the proximity of the proteins used to construct the PAC to the outcome. In other words, a PAC, constructed using proteomic data collected closer to the time of CSVD measurement, was more strongly associated with markers of CSVD. This was further confirmed by testing the associations between PACs using protein selections and weights from different life stages and MRI markers. Midlife PACs, when created using protein weights and selections from the late-life stage, did not show a stronger association with CSVD compared to the original midlife PAC. This suggests that the stronger association between late-life PAC and MRI markers of CSVD, compared to midlife PAC, was not driven by protein weights or selection, but by the timing of protein measurement.

Our findings showed that, in ARIC late-life, higher PAA was more strongly associated with subcortical and lacunar infarcts than with cortical infarcts. Although the biological mechanisms underlying these brain abnormalities remain unclear, a plausible explanation for the differential associations may relate to the anatomical location and characteristics of the affected vessels.

Subcortical and lacunar infarcts are primarily caused by damage to the perforating arteries, which supply blood to the deep regions of the brain (50). These arteries branch directly from the major cerebral arteries, are shorter and narrower than larger cortical surface vessels, and each perforating artery supplies blood to a specific deep-brain territory with very little overlap or collateral connection (51). These characteristics make them more prone to arterial narrowing or occlusion due to cardiovascular risk factors such as hypertension and atherosclerosis, which are common in older adults. As a result, these vessels are more likely to be affected by age-related damage and to develop subcortical or lacunar infarcts, whereas larger cortical vessels are less vulnerable to age-related risk factors (50, 52). Thus, the stronger association observed between late-life PAA and subcortical and lacunar infarcts, compared with cortical infarcts, may suggest that infarcts in these deep brain regions are more strongly affected by accelerated biological aging reflected in PAC.

Also, in ARIC late-life, sensitivity analyses showed that PAA was associated with both strictly lobar and mixed microbleeds, with stronger associations for mixed microbleeds. Strictly lobar microbleeds are primarily attributed to cerebral amyloid angiopathy, a neurodegenerative pathology characterized by amyloid deposition in cortical blood vessels in the lobar regions (53). Mixed microbleeds (the coexistence of microbleeds in both lobar and deep brain regions) involve both cerebral amyloid angiopathy and hypertensive angiopathy (54). Hypertensive angiopathy, largely driven by aging and cardiovascular burden, is a vascular pathology that damages deep-penetrating brain vessels (3, 54). Taken together, our findings indicate that our PAC reflects both cerebral amyloid angiopathy and hypertensive angiopathy, which often coexist in the aging brain and contribute to cognitive impairment.

Our study has several strengths, including a large sample size, the use of proteomic data at different life stages, the use of multiple MRI markers for CSVD, and detailed information on patient characteristics and potential confounders. Also, the relationship between PAC and WMH volume was validated in the external cohort with a broader age range and greater diversity in race/ethnicity.

However, the study also has limitations. First, given the observational design, the results may be influenced by unmeasured or residual confounding. Second, because our study was based on a subsample of ARIC and MESA participants who were invited to and participated in MRI scanning, this may have led to selection bias and limited generalizability. However, in ARIC, we applied inverse probability weighting to account for selection into the brain MRI study. Third, due to the cross-sectional nature of the association between late-life PAA and MRI markers in ARIC, temporality of the association cannot be established. However, it should be highlighted that, in ARIC, we found an association between midlife PAA and WMH volume and subcortical infarcts, assessed nearly 20 years after the protein measurement. Similarly, in MESA, late-life PAA was associated with WMH volume, which was measured approximately eight years after the protein assessment.

Our findings suggest the potential of PACs measured in the early aging stage in predicting later-life small-vessel brain abnormalities, which would enable early risk stratification for more severe clinical outcomes such as increased risk of stroke and dementia even before any symptoms appear (55).

## Data Availability

The data underlying this article will be available upon request and with appropriate approvals

## Authorship Criteria and Responsibilities

S.P., A.E.P., and S.S. conceived and designed the study. S.S., A.E.P., T.M.H., J.C., J.S.P., R.D., J.I.R., P.G., and P.L.L. obtained funding. S.P., S.W., and J.L. conducted the statistical analysis. S.P. wrote the first draft of the manuscript. S.W., T.M.H., E.P.R., J.V., M.H., R.D., R.D., W.S.P., J.I.R., A.C.W., P.G., B.S., W.T., J.C., J.S.P., K.A.W., P.L.L., W.G., A.E.P., and S.S. critically reviewed the results and provided feedback. S.S., A.E.P., and W.G. supervised the study. All authors contributed to revision of the manuscript and approved the final version as submitted.

## Non-standard Abbreviations and Acronyms

ARIC: Atherosclerosis Risk in Communities
CKD-EPI: Chronic Kidney Disease Epidemiology Collaboration
CSVD: cerebral small vessel disease
MESA: Multi-Ethnic Study of Atherosclerosis
MP-RAGE: magnetization-prepared rapid gradient-echo
NCS: Neurocognitive Study
PAC: proteomic aging clock
PAA: proteomic age acceleration
RFU: relative fluorescence units
T2*GRE: T2-weighted gradient-recalled echo
WMH: white matter hyperintensity

## Acknowledgements

The authors thank the other investigators, the staff, and the participants of the ARIC and MESA studies for their valuable contributions.

## Supplemental Material

Tables S1–S7

Figure S1-S8

